# The Prevalence of Neck Pain and Its Association with Studying Device Usage and Posture Among Students at the University of Jordan: A Cross-Sectional Study

**DOI:** 10.1101/2025.06.02.25328842

**Authors:** Mohammad Al-Shalalfeh, Yazan Zayat, Malak Abu Al Haj, Omar Nached, Mohammad Suleiman, Ahmad alaudat, Ahmed Salman

**Author notes:** Ahmed Salman,. These authors contributed equally to this work (Omar Nached, Mohammad Suleiman).

## Abstract

**Background:** Neck pain is one of the leading causes of discomfort and disability, especially among students who use digital devices and books for studying. This study investigates the prevalence of neck pain among students of the University of Jordan and the effects of possible risk factors, such as posture and study utilities.

**Methods:** The data was collected through face-to-face interviews with 507 students stratified across 20 faculties in correspondence to the number of students per faculty. The questionnaire used in the study was structured and assessed demographic factors, study-related behaviors, books and digital device usage, study duration, and physical activity.

**Results:** The data analyzed showed that out of the 507 students, (52.4%) complained of neck pain in the last week, with the most complained site of pain being the neck (56%), and the least being the left shoulder (5.2%). Sitting slouched increased the odds of developing neck pain by 118% compared to sitting upright with full back support (OR = 2.18, CI [1.18, 4.04], p = 0.013). Using laptops (OR = 2.65, CI [1.45, 4.87], p = 0.002) or tablets (OR = 2.68, CI [1.34, 5.36], p = 0.005) for studying was associated with a nearly 2.7-fold increase in the odds of neck pain compared with using phones. In contrast, studying with books was not significantly associated with neck pain. Also, there was no significant difference in odds between tablet and laptop users.

**Conclusion:** This study identified the urgent need for ergonomic education and interventions to promote healthier study habits and reduce musculoskeletal strain in students. Slouched sitting and use of laptops or tablets are risk-increasing, while phone use is risk-decreasing for neck pain development.

## Introduction

Neck pain is a major musculoskeletal disorder and a growing concern in modern society that contributes to disability, economic burden, and reduced quality of life [1].

A global study published in 2021 found that neck pain remains one of the leading causes of years lived with disability (YLDs) worldwide. The study reported that in 2020, neck pain affected 203 million people, with age-standardized prevalence rates being higher in females in comparison to males. By 2050, the number of global neck pain cases is projected to increase by 32.5%, accounting for approximately 269 million cases due to Population growth being the primary contributing factor, followed by population aging [2].

Neck pain, typically associated with occupational settings, especially among those who engage in prolonged desk work, is now an emerging concern among university students. The shift toward digital learning, combined with increased sedentary study habits, has led to an increase in neck pain incidence, with studies reporting rates ranging between 48% and 78% among university students, compared to a general population prevalence rate of 23.1% [3–5].

Among the key risk factors for neck pain in students are sedentary behavior, prolonged screen time, and improper posture, all of which have been exacerbated by the growing dependence on digital devices for daily and academic activities [6].

Prolonged use of mobile phones, laptops, and tablets often results in forward head posture and excessive cervical spine flexion, leading to chronic musculoskeletal discomfort [7].

Furthermore, an observational study has shown that 91% of a sample of 859 students practice a flexed neck posture while using mobile devices, emphasizing a strong correlation between screen exposure and neck pain [8]. Besides, a study by Maayah et al. reported that excessive mobile phone usage, combined with improper posture, is a key contributor to chronic neck and shoulder pain. [9]

Research suggests that maintaining a neutral spine and minimizing static postures may mitigate the progression of neck pain. However, many students engage in improper study practices, such as sitting slouched forward, reading while in bed, or using chairs without back or neck support, all of which increase the strain on the cervical spine, shoulders, and upper extremities, further progressing musculoskeletal discomfort [10–12].

Despite extensive studies done on digital device usage, limited studies have investigated the specific musculoskeletal impacts of various studying utilities, particularly in relation to posture, duration, and individual differences such as sex, which may play a significant role in neck pain development. With academic institutions including both digital and traditional learning methods, it is important to examine how these study methods affect musculoskeletal health. In addition, after reviewing the literature, we found that there is limited literature that identifies the most ergonomically best utilities and studying positions that minimize neck pain, particularly when comparing various digital devices and traditional books. By investigating the association of both digital and book-based studying with neck pain, it will provide a better understanding of risk factors that contribute to neck pain in students.

This study aims to evaluate the prevalence of neck pain among students at the University of Jordan and to examine the association between individual demographic characteristics, sex, history of neck pain, and postural habits while studying including the use of books and digital devices such as (phones, tablets and laptops) with neck pain severity and duration.

## Methods

### 1. Study Design and population

This study applied a cross-sectional design using a structured questionnaire to assess the prevalence, risk factors, and impact of neck pain among university students. The study was carried out at the University of Jordan between 22 December 2024, and 27 January 2025, through face-to-face interviews including students from all 20 faculties at the University of Jordan regardless of their age, sex, or ethnicity. In contrast, students with musculoskeletal diseases, anomalies, or a history of trauma or surgeries in the neck or shoulder regions were excluded from this study.

### 2. Sampling method

A stratified random sampling method was utilized to ensure proportional representation across faculties, with each strata compromising a percentage of our sample in correspondence to the number of students in that faculty. Then, the researchers randomly interviewed students from each faculty.

### 3. Questionnaire Design

The questionnaire covered demographic factors (age, sex, faculty of study), study-related behaviors such as postural habits while studying, books and digital device usage, study duration, and physical activity. To evaluate neck pain, participants reported details about its onset, pain site and duration throughout the week, and intensity using the Numeric Rating Scale (NRS-11), where students were asked to rate their pain on a scale from 0 to 10, where 0 represents “no pain” and 10 represents “worst pain imaginable” [13]. A score rating of 4 was adopted as the threshold for pain severity, as employed by the study Al-Hadidi et al. which followed a similar methodology [14].

Additionally, participants were asked about their approaches for managing this pain, including the use of medication and non-medication interventions such as the usage of heat/cold pads, physical therapy or exercise, and behavioral modifications like adjusting posture or reducing screen time.

Participants were also asked about their knowledge regarding the relationship between posture and neck pain to evaluate participants’ awareness of how ergonomic factors impact musculoskeletal health.

### 4. Ethical approval

The study received ethical approval from the Institutional Review Board (IRB) of the University of Jordan on December 12, 2024 (Decision No. 318/2024), which made certain that every research activity carried out complied with the ethical frameworks and standards. Participants were fully informed about the purpose of this study, and a verbal informed consent was obtained before participation, which was documented through a required confirmation checkbox at the beginning of the online questionnaire. As no identifiable data were collected, responses were confidentially recorded and securely stored to maintain data integrity.

### 5. Statistical analysis

The statistical software JASP (version 0.19.0.0) was used for data processing and analysis. Descriptive statistics, including frequency and percentage, were calculated for categorical variables, while mean and standard deviation were reported for continuous variables.

Univariate and multivariate logistic regression was conducted to estimate the effect of the type of studying utility used on neck pain and pain site and the effect of posture on neck pain utilizing the forced entry selection method. In the model used to estimate the effect of study utilities on neck pain, both shoulders, right shoulder and left shoulder categories were grouped into one category named “shoulder pain”. The results were reported as odds ratio and their 95% confidence interval and a p-value of less than 0.05 was considered statistically significant.

## Results

### Sociodemographic characteristics of the participants

A total of 556 students were asked to participate in the study, with a 91.2% response rate, resulting in 507 valid responses. Our sample was composed of 266 males and 241 females, aged 18 - 29 years old, with a mean age of 21 and a standard deviation of 1.93 years, from different faculties across the University of Jordan. The majority of students (67.2%) spent more than 5 hours a day on their phones for non-studying activities, while (53.6%) spent 3 hours or less a day on studying.

The most used primary utilities for studying were laptops (31.4%) followed by tablets (28.4%), and the least were phones (20.3%) and books (19.9%). On the other hand, phones were the most used secondary devices for studying (31.2%). Most of the students were either sitting slouched (37.3%) or with partial back support (27.8%), while (16%) were sitting with full back support and (3%) without any back support. Tables 1 and 2

**Table 1.** Sociodemographic characteristics:

| Factor | Total,<br>N = 507* | Males,<br>N = 266* | Females,<br>N = 241* |
| --- | --- | --- | --- |
| Age | 21 ± 1.93 | 21.27 ± 2.11 | 20.61 ± 1.64 |
| Faculty |  |  |  |
| School of Medicine | 61 (12%) | 26 (42.6%) | 35 (57.4%) |
| School of Engineering | 51 (10.1%) | 38 (74.5%) | 13 (25.5%) |
| School of Foreign Languages | 49 (9.7%) | 31 (63.3%) | 18 (36.7%) |
| School of Business | 44 (8.7%) | 30 (68.2%) | 14 (31.8%) |
| School of Arts | 37 (7.3%) | 18 (48.6%) | 17 (51.4%) |
| School of Pharmacy | 28 (5.5%) | 6 (21.4%) | 22 (78.6%) |
| School of Science | 27 (5.3%) | 15 (55.6%) | 12 (44.4%) |
| King Abdullah II School of<br>Information Technology | 27 (5.3%) | 21 (77.8%) | 6 (22.2%) |
| School of Agriculture | 26 (5.1%) | 12 (46.2%) | 14 (53.8%) |
| School of Dentistry | 26 (5.1%) | 13 (50%) | 13 (50%) |
| School of Shari'a | 20 (3.9%) | 9 (45%) | 11 (55%) |
| School of Educational<br>Sciences | 19 (3.7%) | 5 (26.3%) | 14 (73.7%) |
| School of Sport Science | 15 (3%) | 8 (53.3%) | 7 (47.7%) |
| School of Rehabilitation<br>Sciences | 14 (2.8%) | 6 (42.9%) | 8 (57.1%) |
| School of Law | 14 (2.8%) | 4 (28.6%) | 10 (71.4%) |
| School of Nursing | 13 (2.6%) | 3 (23.1%) | 10 (76.9%) |
| School of Graduate Studies | 12 (2.4%) | 4 (33.3%) | 8 (66.6%) |
| School of Archaeology and<br>Tourism | 10 (2%) | 7 (70%) | 3 (30%) |
| School of Arts and Design | 9 (1.8%) | 5 (55.6%) | 4 (44.4%) |
| Prince Al Hussein Bin<br>Abdullah II School of<br>International Studies | 5 (1%) | 5 (100%) | 0 (0%) |
| Age | 21 ± 1.93 | 21.27 ± 2.11 | 20.61 ± 1.64 |
| Faculty |  |  |  |
| School of Foreign Languages | 49 (9.7%) | 31 (63.3%) | 18 (36.7%) |
| School of Business | 44 (8.7%) | 30 (68.2%) | 14 (31.8%) |
| School of Arts | 37 (7.3%) | 18 (48.6%) | 17 (51.4%) |
| School of Pharmacy | 28 (5.5%) | 6 (21.4%) | 22 (78.6%) |
| School of Science | 27 (5.3%) | 15 (55.6%) | 12 (44.4%) |
| King Abdullah II School of Information Technology | 27 (5.3%) | 21 (77.8%) | 6 (22.2%) |
| School of Agriculture | 26 (5.1%) | 12 (46.2%) | 14 (53.8%) |
| School of Dentistry | 26 (5.1%) | 13 (50%) | 13 (50%) |
| School of Shari'a | 20 (3.9%) | 9 (45%) | 11 (55%) |
| School of Educational Sciences | 19 (3.7%) | 5 (26.3%) | 14 (73.7%) |
| School of Sport Science | 15 (3%) | 8 (53.3%) | 7 (47.7%) |
| School of Rehabilitation Sciences | 14 (2.8%) | 6 (42.9%) | 8 (57.1%) |
| School of Law | 14 (2.8%) | 4 (28.6%) | 10 (71.4%) |
| School of Nursing | 13 (2.6%) | 3 (23.1%) | 10 (76.9%) |
| School of Graduate Studies | 12 (2.4%) | 4 (33.3%) | 8 (66.6%) |
| School of Archaeology and<br>Tourism | 10 (2%) | 7 (70%) | 3 (30%) |
| School of Arts and Design | 9 (1.8%) | 5 (55.6%) | 4 (44.4%) |
| Prince Al Hussein Bin<br>Abdullah II School of<br>International Studies | 5 (1%) | 5 (100%) | 0 (0%) |

**Table 2.** Device usage and posture.

|  | n (%) |
| --- | --- |
| Hours per day spent on phone |  |
| Less than 1 hour | 2 (0.4%) |
| 1-3 hours | 47 (9.3%) |
| 3-5 hours | 117 (23.1%) |
| 5-7 hours | 166 (32.7%) |
| More than 7 hours | 175 (34.5%) |
| Study duration |  |
| Less than 1 hour | 89 (17.6%) |
| 1-3 hours | 183 (36%) |
| 3-5 hours | 142 (28%) |
| 5-7 hours | 66 (13%) |
| More than 7 hours | 27 (5.3%) |
| Primary studying device |  |
| Tablet | 144 (28.4%) |
| Phone | 103 (20.3%) |
| Laptop | 159 (31.4%) |
| Books | 101 (19.9%) |
| Secondary studying device |  |
| Tablet | 29 (5.7%) |
| Phone | 158 (31.2%) |
| Laptop | 87 (17.1%) |
| Books | 74 (14.6%) |
| None | 159 (31.4%) |
| Posture when studying |  |
| Sitting with full back support | 81 (16%) |
| Sitting slouched | 189 (37.3%) |
| Sitting with partial back support | 141 (27.8%) |
| Sitting without back support | 15 (3%) |
| Lying down | 62 (12.2%) |
| Walking | 19 (3.7%) |

### Prevalence of neck pain and other risk factors

Of the 507 students included in our study, (52.4%) reported neck pain, with the most common site being the cervical region (56%), followed by both shoulders (30.5%), right shoulder (8.3%), and left shoulder (5.2%) Figure 1. (85.6%) scored 4 or less on NRS-11, while the rest scored more than 4.

(40.6 %) of the 266 students who reported neck pain stated that their pain affected the duration they spent on studying and 198 said that they usually change their posture when experiencing pain, (59.1%) of which reported improvement of pain and (35.8%) reported partial improvement. 84 students with neck pain reported using drugs to treat their pain, and 153 students reported usage of non-pharmaceutical methods. The types of both are shown in Figure 2 and Figure 3. Lastly, (86.4%) of the included students were aware of the relationship between posture and neck pain. Table 3

**Table 3.**
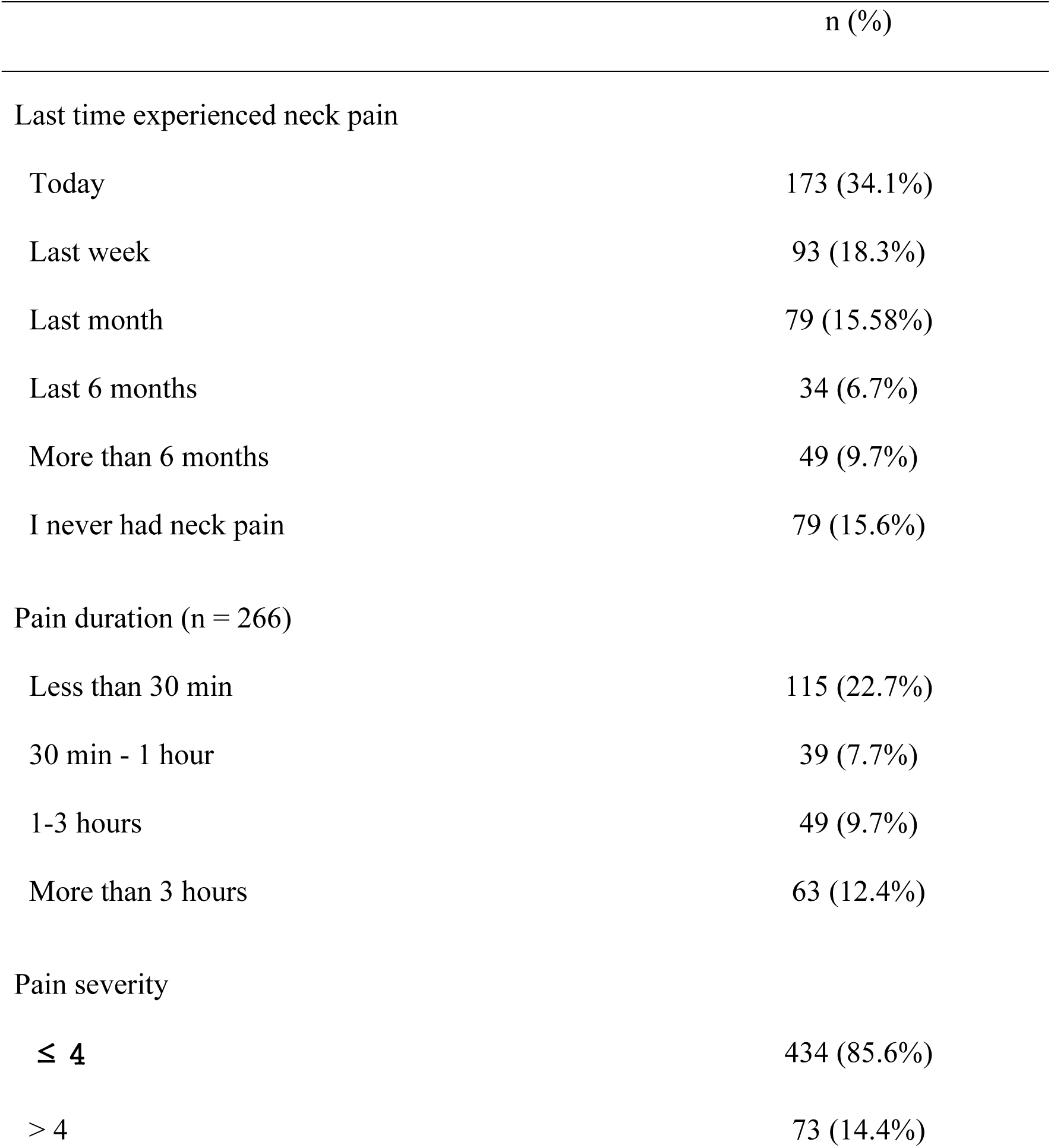
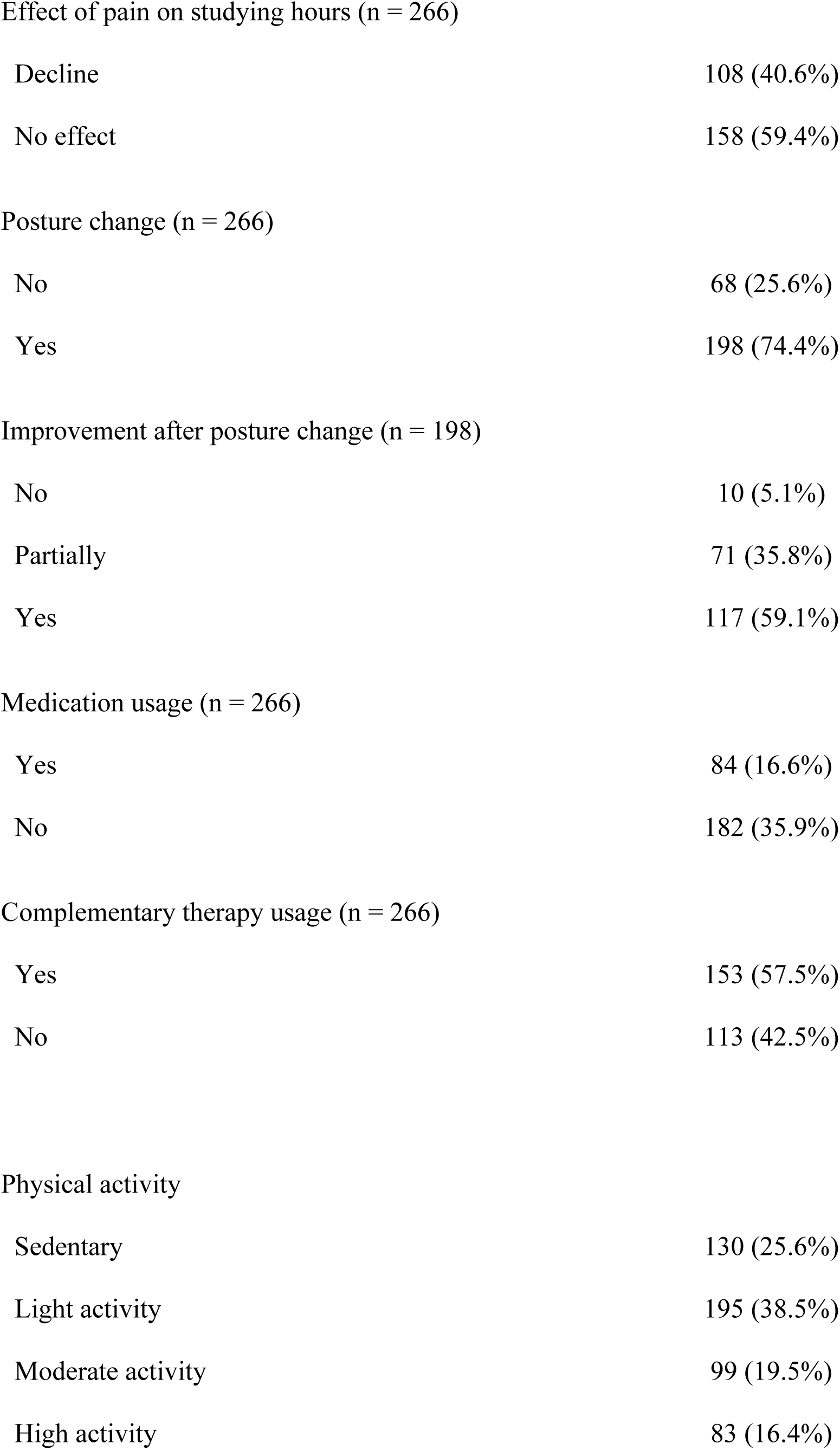
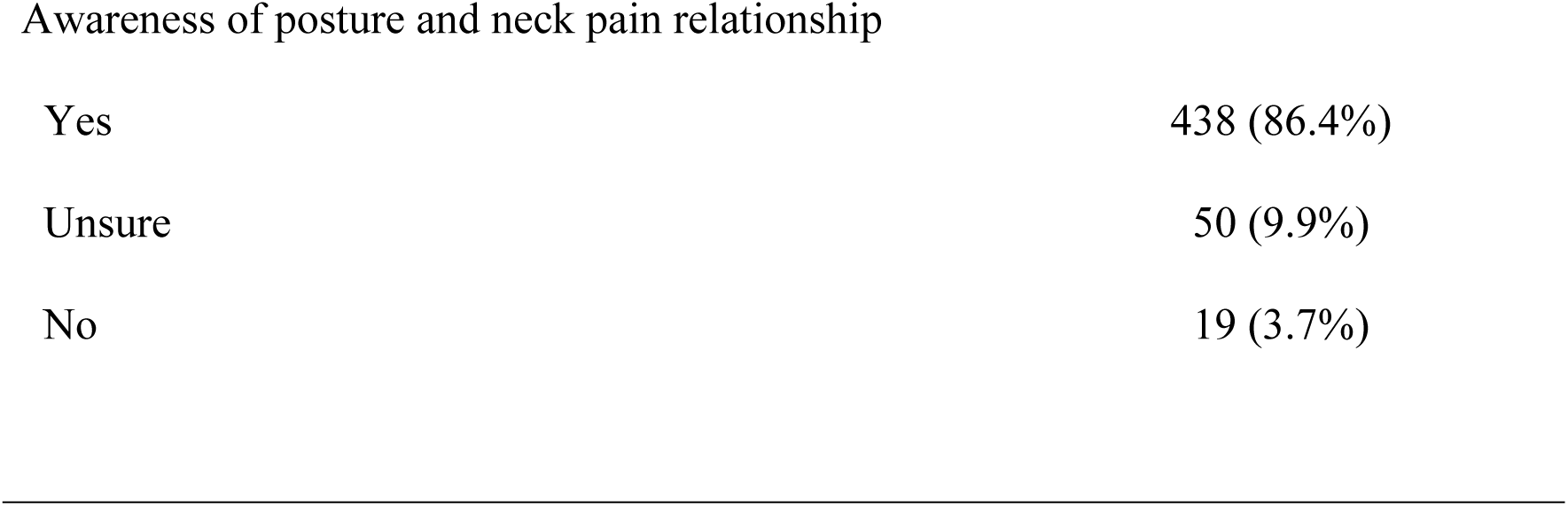
Neck pain prevalence, site, and severity.

|  | n (%) |
| --- | --- |
| Last time experienced neck pain |  |
| Today | 173 (34.1%) |
| Last week | 93 (18.3%) |
| Last month | 79 (15.58%) |
| Last 6 months | 34 (6.7%) |
| More than 6 months | 49 (9.7%) |
| I never had neck pain | 79 (15.6%) |
| Pain duration (n = 266) |  |
| Less than 30 min | 115 (22.7%) |
| 30 min - 1 hour | 39 (7.7%) |
| 1-3 hours | 49 (9.7%) |
| More than 3 hours | 63 (12.4%) |
| Pain severity |  |
| $\leq 4$ | 434 (85.6%) |
| $> 4$ | 73 (14.4%) |
| Decline | 108 (40.6%) |
| No effect | 158 (59.4%) |
| Posture change (n = 266) |  |
| No | 68 (25.6%) |
| Yes | 198 (74.4%) |
| Improvement after posture change (n = 198) |  |
| No | 10 (5.1%) |
| Partially | 71 (35.8%) |
| Yes | 117 (59.1%) |
| Medication usage (n = 266) |  |
| Yes | 84 (16.6%) |
| No | 182 (35.9%) |
| Complementary therapy usage (n = 266) |  |
| Yes | 153 (57.5%) |
| No | 113 (42.5%) |
| Physical activity |  |
| Sedentary | 130 (25.6%) |
| Light activity | 195 (38.5%) |
| Moderate activity | 99 (19.5%) |
| High activity | 83 (16.4%) |
| Yes | 438 (86.4%) |
| Unsure | 50 (9.9%) |
| No | 19 (3.7%) |

### Association between neck pain and other risk factors

In females the odds of experiencing neck pain increased by 2.5 times (OR = 2.52, CI [1.76, 3.6], *p* = < 0.001), sitting slouched doubled the odds (OR = 2.03, CI [1.2, 3.44], *p* = 0.008), and studying for 5-7 hours a day increased the odds by 3 times (OR = 3.08, CI [1.55, 6.15], *p* = 0.001). On the contrary moderate physical activity reduced the odds of developing neck pain (OR = 0.61, CI [0.37, 0.99], *p* = 0.044). Furthermore, using tablets increased the odds for neck pain development by 2.22 times when compared to using phones (OR = 2.22, CI [1.32, 3.71], *p* = 0.002). Interestingly, we found that students at King Abdullah II School of Information Technology faculty had a reduced odds of developing neck pain when compared to the faculty of medicine. Regarding pain site, in females the odds of developing pain at the shoulders was 1.87 times higher when compared to the cervical region (OR = 1.87, CI [1.13, 3.09], *p* = 0.014), while using tablets or laptops reduces the odds by 0.32 times when compared to using books (OR = 2.7, CI [0.13, 0.56], *p* = < 0.001) and (OR = 0.32, CI [0.16, 0.66], p = 0.002), respectively. The rest of the variables were not statistically associated with either neck pain or pain site. Tables 4,5, and 6.

### Multivariate analysis

To further investigate the effect of primary device usage on neck pain and pain site and posture on neck pain, we built a logistic regression model.

Regarding the model used to estimate the effect of study device usage and pain site, we included sex as a potential confounder and physical activity and age as independent predictors of the outcome. We found that in comparison to using books as a study device, using tablets decreases the odds of experiencing cervical pain compared to shoulder pain by approximately 74% (OR = 0.261, CI [0.12, 0.55], p < 0.001). Moreover, using a laptop as a study device decreases the odds of experiencing cervical pain compared to shoulder pain by 60% (OR = 0.4, CI [0.19, 0.85], p = 0.017). Table 4

**Table 4.**
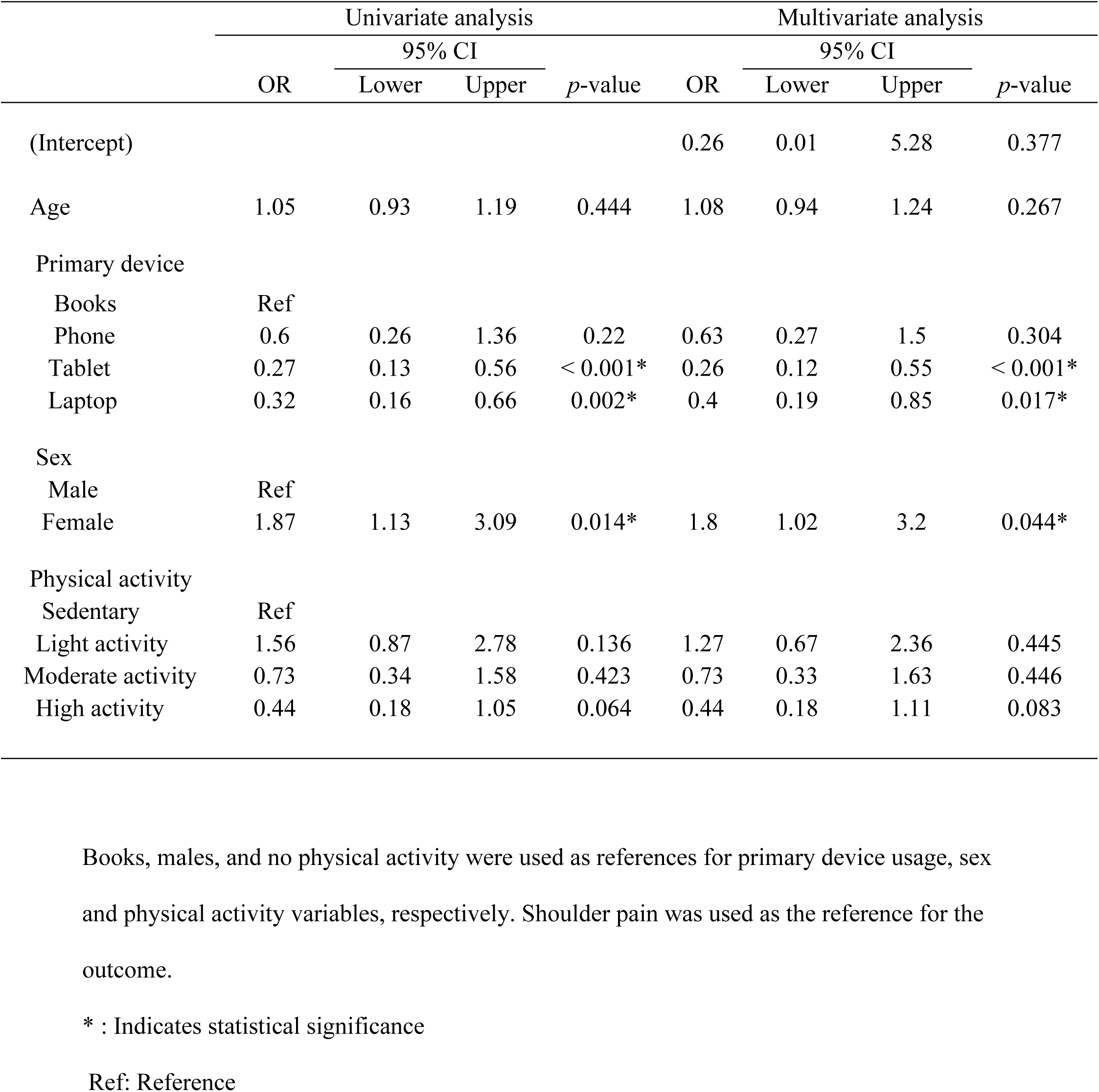
Logistic regression model for the estimate of the effect of device usage on pain.

|  | Univariate analysis |  |  |  | Multivariate analysis |  |  |  |
| --- | --- | --- | --- | --- | --- | --- | --- | --- |
|  | OR | 95% CI |  | *p*-value | OR | 95% CI |  | *p*-value |
|  |  | Lower | Upper |  |  | Lower | Upper |  |
| (Intercept) |  |  |  |  | 0.26 | 0.01 | 5.28 | 0.377 |
| Age | 1.05 | 0.93 | 1.19 | 0.444 | 1.08 | 0.94 | 1.24 | 0.267 |
| Primary device |  |  |  |  |  |  |  |  |
| Books | Ref |  |  |  |  |  |  |  |
| Phone | 0.6 | 0.26 | 1.36 | 0.22 | 0.63 | 0.27 | 1.5 | 0.304 |
| Tablet | 0.27 | 0.13 | 0.56 | < 0.001* | 0.26 | 0.12 | 0.55 | < 0.001* |
| Laptop | 0.32 | 0.16 | 0.66 | 0.002* | 0.4 | 0.19 | 0.85 | 0.017* |
| Sex |  |  |  |  |  |  |  |  |
| Male | Ref |  |  |  |  |  |  |  |
| Female | 1.87 | 1.13 | 3.09 | 0.014* | 1.8 | 1.02 | 3.2 | 0.044* |
| Physical activity |  |  |  |  |  |  |  |  |
| Sedentary | Ref |  |  |  |  |  |  |  |
| Light activity | 1.56 | 0.87 | 2.78 | 0.136 | 1.27 | 0.67 | 2.36 | 0.445 |
| Moderate activity | 0.73 | 0.34 | 1.58 | 0.423 | 0.73 | 0.33 | 1.63 | 0.446 |
| High activity | 0.44 | 0.18 | 1.05 | 0.064 | 0.44 | 0.18 | 1.11 | 0.083 |

Books, males, and no physical activity were used as references for primary device usage, sex
and physical activity variables, respectively. Shoulder pain was used as the reference for the
outcome.
\* : Indicates statistical significance
Ref: Reference

Regarding the model used for study devices and neck pain, we included sex and faculty as potential confounders and age as an independent predictor. We excluded posture and study duration since they were likely to mediate the effect of study devices on neck pain. We also analyzed the effect of primary devices on neck pain 3 times, each time with a different reference to get a better understanding on how devices affect the development of neck pain. Using laptops (OR = 2.65, CI [1.45, 4.87], p = 0.002) or tablets (OR = 2.68, CI [1.34, 5.36], p = 0.005) for studying increased the odds for developing neck pain nearly 2.7 times when compared to using phones. Meanwhile, there wasn’t any statistically significant evidence that studying using books increased or decreased the odds for developing pain when compared with the rest of devices, (tablets (OR = 1.82, CI [0.91, 3.65], p = 0.092), laptops (OR = 1.8, CI [0.97, 3.37], p = 0.064) and phones (OR = 0.68, CI [0.36, 1.27], p = 0.224)), respectively.

Moreover, we did not find a statistically significant change in the odds between tablet users and laptop users. Table 5

**Table 5.** Logistic regression model for the estimate of the effect of device usage on neck pain.

|  | Univariate analysis |  |  |  | Multivariate analysis |  |  |  |
| --- | --- | --- | --- | --- | --- | --- | --- | --- |
|  | OR | 95% CI |  | *p*-value | OR | 95% CI |  | *p*-value |
|  |  | Lower | Upper |  |  | Lower | Upper |  |
| (Intercept) |  |  |  |  | 0.19 | 0.02 | 2.32 | 0.193 |
| Study device |  |  |  |  |  |  |  |  |
| Phone | Ref |  |  |  |  |  |  |  |
| Tablet | 2.22 | 1.32 | 3.71 | 0.002* | 2.68 | 1.34 | 5.36 | 0.005* |
| Laptop | 1.59 | 0.96 | 2.62 | 0.071 | 2.65 | 1.45 | 4.87 | 0.002* |
| Books | 1.67 | 0.96 | 2.91 | 0.07 | 1.47 | 0.79 | 2.75 | 0.224 |
| Study device |  |  |  |  |  |  |  |  |
| Books | Ref |  |  |  |  |  |  |  |
| Tablet | 1.33 | 0.79 | 2.22 | 0.279 | 1.82 | 0.91 | 3.65 | 0.092 |
| Laptop | 0.95 | 0.58 | 1.57 | 0.842 | 1.8 | 0.97 | 3.37 | 0.064 |
| Phone | 0.6 | 0.34 | 1.04 | 0.07 | 0.68 | 0.36 | 1.27 | 0.224 |
| Study device |  |  |  |  |  |  |  |  |
| Tablet | Ref |  |  |  |  |  |  |  |
| Phone | 0.45 | 0.27 | 0.76 | 0.002* | 0.37 | 0.19 | 0.75 | 0.005* |
| Laptop | 0.72 | 0.45 | 1.13 | 0.151 | 0.99 | 0.54 | 1.82 | 0.977 |
| Books | 0.75 | 0.45 | 1.26 | 0.279 | 0.55 | 0.27 | 1.1 | 0.092 |
| Age | 1.02 | 0.93 | 1.12 | 0.642 | 1.06 | 0.94 | 1.19 | 0.328 |
| Sex |  |  |  |  |  |  |  |  |
| Male | Ref |  |  |  |  |  |  |  |
| Female | 2.52 | 1.76 | 3.6 | < 0.001* | 2.44 | 1.62 | 3.7 | < 0.001* |
| Faculty |  |  |  |  |  |  |  |  |
| School of medicine | Ref |  |  |  |  |  |  |  |
| School of Business | 2.07 | 0.92 | 4.66 | 0.077 | 4.05 | 1.58 | 10.36 | 0.004* |
| School of Law | 3.55 | 0.9 | 13.99 | 0.07 | 6.09 | 1.34 | 27.73 | 0.019* |
| School of Arts and | 3.39 | 0.65 | 17.63 | 0.147 | 6.16 | 1.04 | 36.56 | 0.046* |
| Design |  |  |  |  |  |  |  |  |
| Physical activity |  |  |  |  |  |  |  |  |
| Sedentary | Ref |  |  |  |  |  |  |  |
| Light activity | 1.36 | 0.86 | 2.14 | 0.184 | - | - | - | - |
| Moderate activity | 0.61 | 0.37 | 0.99 | 0.044* | - | - | - | - |
| High activity | 0.63 | 0.38 | 1.06 | 0.08 | - | - | - | - |

Tablets, males, and the school of medicine were used as references for primary device usage, sex, and faculty variables, respectively. The absence of neck pain was used as the reference for the outcome. Only the faculties that were associated with neck pain are presented in the table, detailed results can be found in the supplementary files.
\*: Indicates statistical significance
Ref: Reference

The model used for estimating the effect of posture on neck pain included sex, faculty, primary device usage, and study hours as confounders and age as an independent predictor. Sitting slouched increases the odds of developing neck pain by 118% compared to sitting upright with full back support (OR =2.18, CI [1.18, 4.04], p = 0.013). Table 6

**Table 6.**
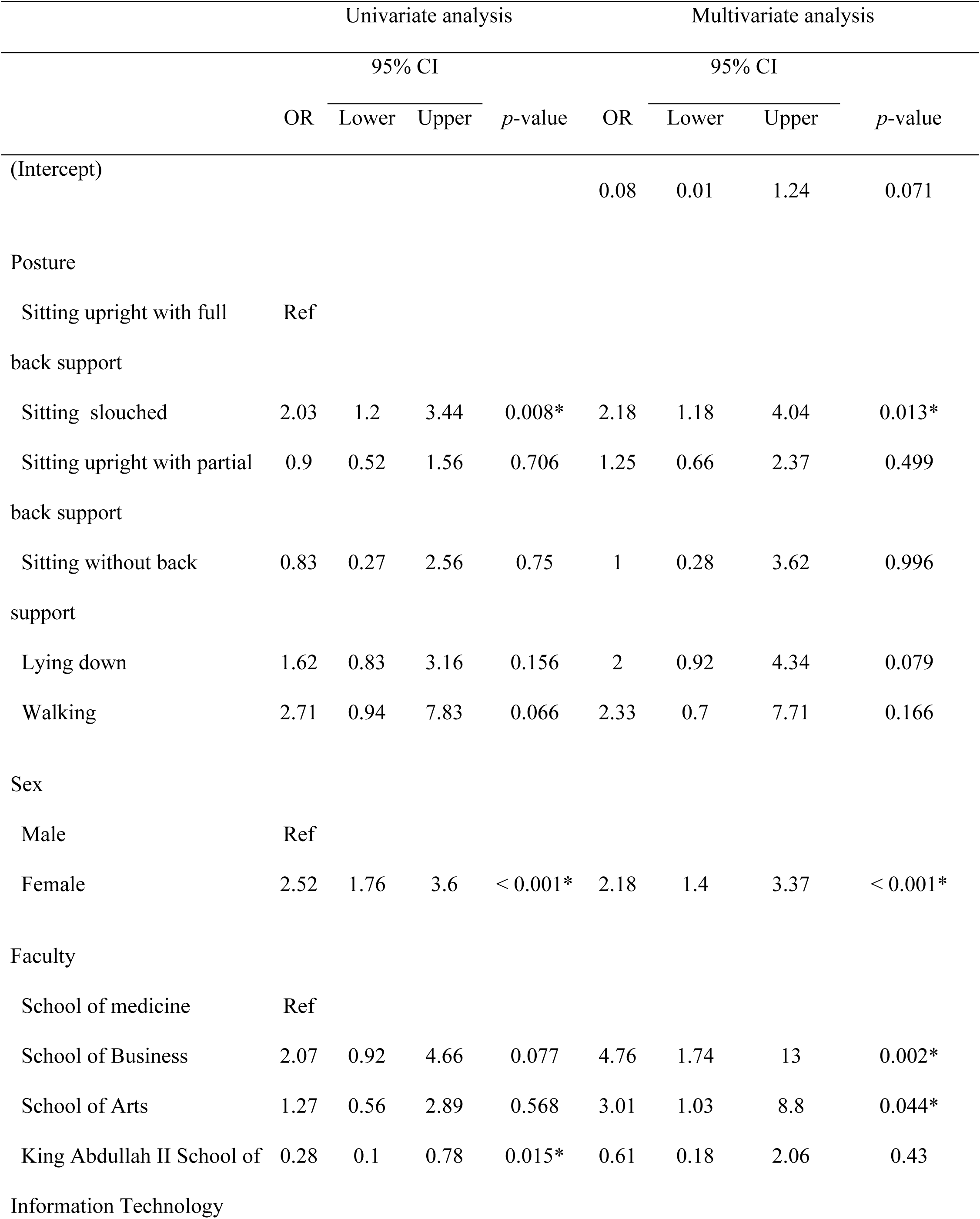
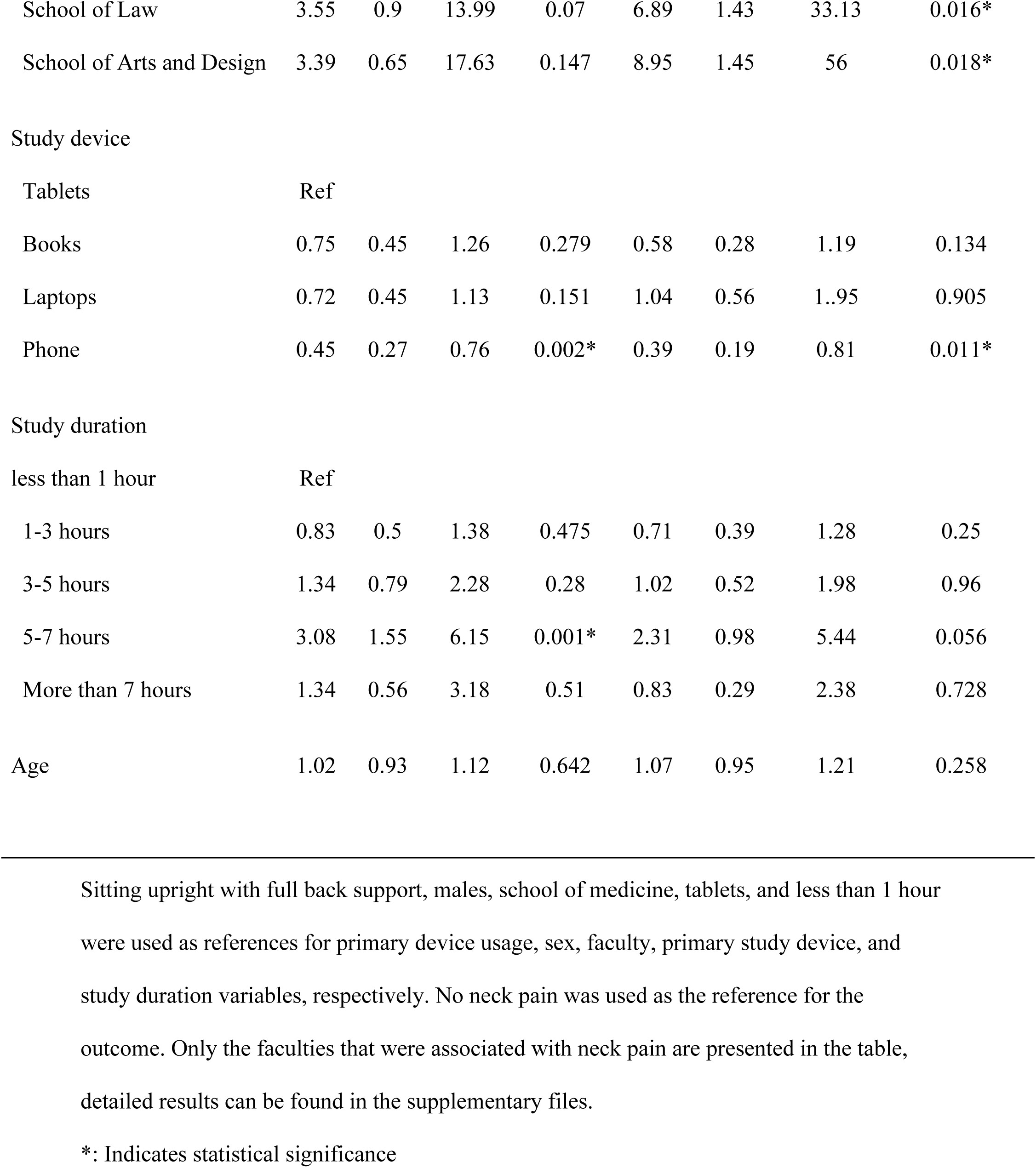
Logistic regression model for the estimate of the effect of posture on neck pain.

## Discussion

In this study, our sample revealed that most of the students at the University of Jordan have experienced neck pain during the week prior to the interview, with several factors showing a statistically significant association. Univariate analysis showed that students who studied for 5-7 hours per day were 3.08 times more likely to experience neck pain compared to those who studied for shorter durations. Female students were 2.52 times as likely to report neck pain as male students. Furthermore, multivariate analysis showed students who studied on tablets increased their odds of experiencing neck pain by 2.68 times compared to those who studied on mobile phones.

Apart from faculty and lack of physical activity being linked with neck pain, sitting in a slouched posture doubled the odds of experiencing neck pain.

All of the aforementioned were statistically significant risk factors for neck pain. Our results show that there is an increasing incidence of neck pain among students, especially after the shift to digital-based learning and work, which became prevalent during the COVID-19 pandemic.

Before this shift, the prevalence of neck pain among university students was reported to be 37.3%, which then increased to 62.7% [15]. This trend is in concordance with previous research that highlighted the musculoskeletal complications that result from prolonged exposure to digital devices. In our sample of 507 students, 52.4% experienced neck pain in the past week, with the cervical region being the most affected region, followed by both shoulders. In addition, we found that laptop and tablet users had a higher prevalence of neck pain than mobile phone users. Moreover, the results of our study also support previous findings showing that females reported higher pain levels than males.

Our findings of neck pain prevalence among university students align with studies that evaluated neck pain in 7 days. For example, a study done among medical students at the University of Ankara, Turkey, reported a 50.6% prevalence of neck pain [16], which closely matches our observed result of 52.4%. Similarly, a Canadian study also found comparable one-week prevalence rates, ranging from 63.7% among faculty of Health sciences students, 45.4% among Education faculty students, to 76.9% among chiropractic students [17].

These variations could be attributed to differences in device usage habits, culture, ergonomic awareness, and educational interventions.

The increasing reliance on digital devices has significantly influenced both work and study environments, requiring users to engage in suboptimal postures during device usage, further increasing their risk of developing neck pain.

A meta-analysis by Gao et al. reported that university students who used digital devices for extended periods had 53% higher odds of developing neck pain. [4]

Our multivariate analysis revealed that tablet and laptop users had nearly 2.7 times greater odds of developing neck pain when compared to phone usage. This makes phones a better studying device to use, in terms of neck pain risk, than tablets and laptops, but not any different from studying from books.

In regards to the site of neck pain, tablet use was also strongly associated with shoulder pain, increasing the likelihood of developing shoulder pain by nearly four times more than neck pain when compared to using books.

These results highlight that mobile users experienced the least musculoskeletal discomfort in this sample. Despite their small screen size and suboptimal ergonomic design, mobile phones turned out to be a better study utility compared to laptops or tablets. One study found that phone users tend to adjust their postures more frequently than laptop and tablet users, reducing static muscle load and strain. Therefore, reducing the risk of neck pain development. [18]

Postural habits were another significant risk factor for neck pain in our study. Our results showed that students who maintained a slouched posture were 118% more likely to develop neck pain compared to those who sat upright with full back support. This result aligns with multiple studies that have linked forward bent posture and prolonged static postures with an increased risk of neck pain occurrence.

A study by Cagnie et al. reported that frequently having the neck in a forward bent posture for prolonged durations increased the odds of developing neck pain by two times, with prolonged sitting further raising the risk by nearly two times as well. [19]

Similarly, a meta-analysis study by Gao et al. further supports these findings, reporting an odds ratio of 2.04 for prolonged head-bowing, contributing to neck pain development. [4]

Overall, these studies emphasize that practicing improper postures, particularly slouched posture, increases cervical spine stress, leading to neck pain. In our sample, we found that female students were more likely to report neck pain than males [21]. This finding is consistent with previous research highlighting sex differences in musculoskeletal complaints. For instance, a study done in a Chinese high school showed that Neck/shoulder pain and low back pain were reported more frequently by females than males [19,20] and according to Salameh et al., among Jordanian students studying medicine, it revealed females are more affected by text neck syndrome than males.

This result could be attributed to the biological differences in nociceptor density. Women are known to have a higher density of nociceptors in the skin and muscles, leading to greater sensitivity to pain stimuli. Furthermore, hormonal fluctuations affect pain perception, with studies indicating that high estrogen levels can increase pain sensitivity as it is shown to have a pro-nociceptive effect. Whereas testosterone has a protective, anti-nociceptive effect [22]. This could explain why women report higher pain intensity than men. Moreover, sociocultural factors also contribute to sex differences in neck pain prevalence. Studies indicate that expressing pain is more socially acceptable among women, leading to higher self-reported neck pain rates compared to men [23].

An interesting finding is that the majority of students (67.2%) spent more than 5 hours per day on their phones without studying, while (53.6%) of students spent 3 hours or less daily on studying. These findings show that many students spend a large amount of time on distractions rather than studying, which may reduce their productivity, competency, and academic responsibility. Consequently, this could impact their ability to acquire essential knowledge and skills for their future careers, potentially affecting the career growth of future generations. Furthermore, these findings raise concerns about the potential risks the newest generation of students are facing from this unhealthy lifestyle, since it has been shown that students who used their smartphones for more than four hours daily had increased odds of experiencing poor sleep, anxiety, stress, and even low academic performance as established by Nikolic et al. [24].

In this study, we investigated the prevalence of neck pain and how specific postures and the use of certain studying utilities contribute to the risk of developing this condition.

Contrary to several studies conducted in the region, our study employed multivariate methods for data analysis, which reduced the degree of bias introduced by confounders. Furthermore, in contrast to a study [14,25] which was conducted at the University of Jordan, specifically the health care faculties, the data utilized in our study was collected through face-to-face interviews, which increased the reliability and accuracy of responses collected. Moreover, we stratified our sample population across all faculties to increase the generalizability of the data.

## Limitations

The study included only University Of Jordan students, limiting the generalizability to other people in Jordan. The cross-sectional design could not determine the direction of causality between these variables. Although we stratified our sample, we still used convenience sampling to collect data from each stratum, resulting in some sampling bias.

## Conclusion

We found that most students had neck pain in the week before the interview. Sitting slouched while studying with common study devices increases the risk of neck and shoulder pain more than two times sitting upright with full back support. Tablet devices increase the risk of neck and shoulder pain by approximately three times more than phones. There was no statistical difference between using books, tablets, and laptops. Additionally, using tablets and laptops as studying utilities tend to cause pain in the shoulder region more than the cervical region when compared to studying from books.

## Recommendations

Sitting positions and studying devices are associated positively with neck pain, which is a rising problem among students. More attention should be given to increasing awareness about proper sitting postures to reduce the prevalence of neck and shoulder pain in society.

## Data Availability

The collected data may be requested from the corresponding author.

https://drive.google.com/drive/folders/17zDLTQRNkjsKRtj5Z6a7jauwxI-KY2mH?usp=sharing

